# Enhanced mindfulness based stress reduction (MBSR+) in episodic migraine: a randomized clinical trial with MRI outcomes

**DOI:** 10.1101/19004069

**Authors:** David A. Seminowicz, Shana AB Burrowes, Alexandra Kearson, Jing Zhang, Samuel R Krimmel, Luma Samawi, Andrew J Furman, Michael L Keaser, Neda F. Gould, Trish Magyari, Linda White, Olga Goloubeva, Madhav Goyal, B. Lee Peterlin, Jennifer A. Haythornthwaite

## Abstract

We aimed to evaluate the efficacy of an enhanced mindfulness based stress reduction (MBSR+) versus stress management for headache (SMH). We performed a randomized, assessor-blind, clinical trial of 98 adults with episodic migraine recruited at a single academic center comparing MBSR+ (n=50) to SMH (n=48). MBSR+ and SMH were delivered weekly by group for 8 weeks, then bi-weekly for another 8 weeks. The primary clinical outcome was reduction in headache days from baseline to 20 weeks. MRI outcomes included activity of left dorsolateral prefrontal cortex (DLPFC) and cognitive task network during cognitive challenge, resting state connectivity of right dorsal anterior insula (daINS) to DLPFC and cognitive task network, and gray matter volume of DLPFC, daINS, and anterior midcingulate. Secondary outcomes were headache-related disability, pain severity, response to treatment, migraine days, and MRI whole-brain analyses. Reduction in headache days from baseline to 20 weeks was greater for MBSR+ (7.8 [95%CI, 6.9-8.8] to 4.6 [95%CI, 3.7-5.6]) than for SMH (7.7 [95%CI 6.7-8.7] to 6.0 [95%CI, 4.9-7.0]) (*P*=0.04). 52% of the MBSR+ group showed a response to treatment (50% reduction in headache days) compared with 23% in the SMH group (*P*=0.004). Reduction in headache-related disability was greater for MBSR+ (59.6 [95%CI, 57.9-61.3] to 54.6 [95%CI, 52.9-56.4]) than SMH (59.6 [95%CI, 57.7-61.5] to 57.5 [95%CI, 55.5-59.4]) *(P=*0.02). There were no differences in clinical outcomes at 52 weeks or MRI outcomes at 20 weeks, although changes related to cognitive networks with MBSR+ were observed. MBSR+ is an effective treatment option for episodic migraine.

## Introduction

Migraine is a severe and often disabling neurological disorder [21:44] and standard preventative agents frequently create challenging side-effects [49:57]. Migraine guidelines [51] include nonpharmacological preventative treatments, and mindfulness-based stress reduction (MBSR) recently has been shown to improve pain and functional outcomes in chronic low back pain [19]. Yet meditation and mindfulness therapies show only modest benefits to date in reducing the frequency of migraine [4:35:76]. The outcomes of MBSR in reducing pain [33] and migraine frequency [4] may improve if training is enhanced to include a longer period of learning, as greater home practice yields better outcomes in MBSR [50]. Since medication can contribute to the frequency of headache [14], MBSR may be an effective nonpharmacological prevention strategy that has become widely available throughout the United States and Europe in recent years.

Migraine headaches are due to acute alterations in the trigeminovascular system and changes in brain perfusion include widespread increases and decreases in brain activity [13:18:22]. Beyond the changes known to occur during attacks, mild cognitive deficits occur between attacks [71:75] and brain structure is altered relative to controls [8:39]. These brain changes involve cognitive and emotional circuits [20], particulary the insula [11], left dorsolateral prefrontal cortex (DLPFC), and anterior/mid cingulate cortex (ACC/MCC) [48]. Our previous work on pain and cognition has focused on the role of the DLPFC and the cognitive task network known as the extrinsic mode network (EMN) – which is a brain network activated across multiple types of cognitive challenges (e.g. conflict, working memory) and anti-correlated with the default mode network [38] – and the connectivity of these regions to the anterior insula[16:58:60:61:63:66]. The demands of recurring pain deplete cognitive and emotional resources [58], and treatments that increase the efficiency of information processing, or cognitive efficiency, may be particularly beneficial for painful conditions such as migraine. While mindfulness meditation can reduce acute pain through complex cortical and thalamic mechanisms that are independent of endogenous opioids[80-84], long-term practice appears to increase cognitive efficiency. Long-term meditation practitioners show structural changes in brain areas involved in cognitive and emotional processing (insula, ACC/MCC, and prefrontal cortex [28]), and mindfulness training changes brain function in these and other areas, with consistent long-term changes in insula cortex [32:79]. The focused attention involved in mindfulness activates these cognitive networks [23] and even brief mindfulness training improves cognitive efficiency and increases engagement of left DLPFC [3]. Increased cognitive efficiency contributes to control over pain in long-term mindfulness practitioners [34].

This trial compared enhanced MBSR (MBSR+) to an active control on clinical and imaging outcomes in episodic migraine. We hypothesized that MBSR+ would reduce headache frequency (primary) and reduce migraine-related disability (secondary). We also hypothesized that MBSR+ would alter the structure and function of brain areas and networks involved in cognitive efficiency, including: increased gray matter volume in the DLPFC, MCC, and insula; decreased activation of left DLPFC and EMN during cognitive challenge; and reduced resting state connectivity from anterior insula to left DLPFC and cognitive task network.

## Methods

The study protocol was approved by the Johns Hopkins School of Medicine and the University of Maryland Baltimore Institutional Review Boards. Participants were recruited from local headache clinics, primary care providers, and the community in eight cohorts (9-18 participants/cohort) from June 2014 to February 2017. Cohorts included participants randomized to SMH or MBSR+, with both study arms running concurrently. Recruited individuals were 18 to 65 years of age and met International Classification of Headache Disorders criteria for migraine with or without aura [1]. Eligibility was assessed first by telephone (Figure 1), then a screening visit. Following written informed consent, screening established >1 year history of a migraine diagnosis and excluded individuals who reported severe or unstable psychiatric symptoms, used opioid medications, had prior experience with mindfulness or concurrent treatment expected to affect mindfulness/stress reduction (see Protocol for full inclusion/exclusion). Potential participants completed at least 28 days of an electronic daily diary to establish eligibility (4-14 headache days in 28 days) which served as the baseline measure of headache frequency. Eligible subjects then attended the MRI session, including written informed consent, questionnaires, and quantitative sensory testing.

**Figure 1.**
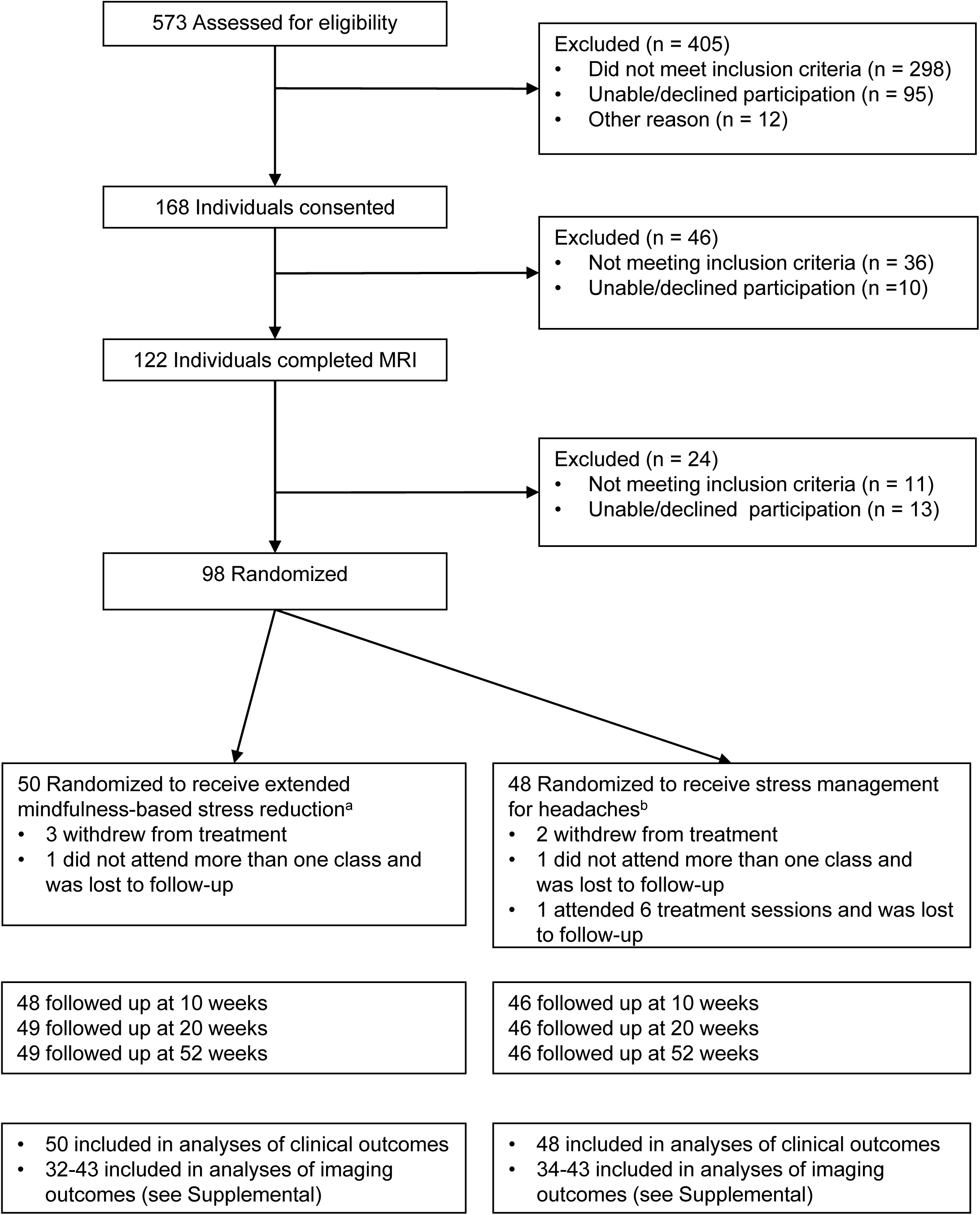
Participant Flow Through Trial Comparing Extended Minfulness Based Stress Reduction and Stress Management for Migraine Headache. ^a^ – Of the 50 participants randomized to receive extended mindfulness-based stress reduction, 43 completed all 12 sessions. Participants were recruited in 8 separate cohorts (range of 4-8/cohort). ^b^ – Of the 48 participants randomized to receive stress management for headaches, 40 completed all 12 sessions. Participants were recruited in 8 separate cohorts (range of 2-11/cohort).

### Assessments

Study questionnaires were completed online at baseline, week 10 (after 8-weeks of MBSR+/SMH), week 20 (after completion of MBSR+/SMH), and at week 52. MRI visits at baseline, week 10, and week 20 were conducted by staff masked to treatment group. All MRI scans used a Siemens Tim-Trio 3T MRI scanner with a 32-channel head coil through March 2017, then a Siemens 3T Prisma Fit MRI scanner with a 64-channel head coil (see below for details). Scans included a T1 high resolution anatomical scan, a resting state fMRI scan, and fMRI scans during completion of two blocks of painful thermal stimulation and two blocks of cognitive challenge (the multi-source interference task (MSIT) [15].

### Randomization

Randomization (1:1) was stratified by the presence/absence of another chronic pain disorder and headache frequency from the baseline headache diary (low: 4-8; high: 9-14 headache days per 28 days). The randomization schedule was generated online (randomization.com) and stored in a locked cabinet by non-study staff. Assignment occurred by non-study staff when the participant arrived for the first day of class. Thus, intervention staff were blinded to assignment until the first day of class. Participants were instructed not to discuss their assignment with assessors, who were blinded to assignment. The principal investigator and statistician remained blinded until completion of the study and analysis.

### Interventions

Participants were instructed to continue stable use of prescribed preventative treatments and continue use of acute abortives as needed. Separate groups for each intervention met for about 2 hours weekly for 8 weeks then bi-weekly for another 8 weeks. A trained expert in the content for each intervention used a manualized protocol that included participant handouts and materials for home use. MBSR+ was administered by 2 experienced, certified instructors (10 and 40 years of meditation experience). SMH was delivered by a nurse practitioner (11 years experience treating headache patients). Checklists were completed by instructors at the end of sessions to verify all components were delivered. Missed sessions were made up individually in person or by phone.

The enhanced MBSR (MBSR+) included 12 sessions over 4 months, including eight weekly sessions followed by four bi-weekly sessions. The first eight sessions adapted the MBSR program developed by Jon Kabat-Zinn [40] to include trauma-informed methods of teaching and emphasized loving kindness to distress [42]. Study participants were provided with audio CDs and handouts and a personal copy of *Full Catastrophe Living* by Jon Kabat-Zinn. Each session included a longer arriving practice, and a loving kindness meditation was included at week 2 and at the retreat, held between weeks 6 and 8. The week 8 class was adapted to focus on applying learning to migraines before, during and after an attack and engaging participants in deciding which MBSR practices they wished to increase practice of during the second eight weeks of the MBSR+ program. The additional four bi-weekly sessions enhanced typical MBSR training by encouraging continued mindfulness practice and self-compassion and emphasizing sympathetic joy, equanimity, and gratitude. The format of these bi-weekly sessions was similar to the original program and included both didactic content and mindfulness practice, including body scan, yoga, sitting and walking meditations.

SMH included 12 sessions over 4 months focused on didactic content about the role of stress and other triggers in headaches and followed a smiliar format and timing to the MBSR+ sessions, minus the retreat. Topics included stress at work and home; coping with stress mental health and personality; sleep hygiene; pain education; and medications for migraine. Information, group discussion, and social support among group members was emphasized; behavior change and specific skill development was not addressed. Each session included a 10-minute period of standardized muscle stretching exercises. In addition to educational handouts, participants were provided with a personal copy of *The Migraine Brain* by Carolyn Bernstein.

### Measures

Sociodemographic and medical data were obtained at baseline (Table 1). Clinical and imaging outcome measures were collected at baseline, at week 10 after the first phase of MBSR+/SMH (secondary), and week 20 after the second phase of MBSR+/SMH (primary) and clinical outcome measures were collected at week 52 (secondary). The week 20 time point, along with the 28 day period of prospective diaries that patients completed, conforms to current guidelines on RCTs for migraine prophylaxis [69].

**Table 1.** Sample characteristics. N (%) unless otherwise stated.

| Characteristic | All , n=98 | Randomized to SMH, n=48 | Randomized to MBSR+, n=50 | p-value |
| --- | --- | --- | --- | --- |
| Age: median years (range) | 36 (18-65) | 36 (21-63) | 36 (18-65) | NS |
| Gender |  |  |  |  |
| M | 9 (9.2) | 6 (12.5) | 3 (6.0) | NS |
| F | 89 (90.8) | 42 (87.5) | 47 (94.0) |  |
| Race |  |  |  |  |
| White | 71 (72.4) | 36 (75.0) | 35 (70.0) | NS |
| African American | 17 (17.3) | 7 (14.6) | 10 (20.0) |  |
| Other | 9 (9.1) | 5 (10.4) | 4 (8.0) |  |
| Presence of idiopathic pain |  |  |  |  |
| No | 70 (71.4) | 35 (72.9) | 35 (70.0) | NS |
| Yes | 28 (28.6) | 13 (27.1) | 15 (30.0) |  |
| Headache Frequency |  |  |  |  |
| Low (4-8) | 50 (51.0) | 25 (52.1) | 25 (50.0) | NS |
| High (>8 and <15) | 48 (49.0) | 23 (47.9) | 25 (50.0) |  |
| Education |  |  |  |  |
| Up to some college | 20 (20.4) | 7 (14.6) | 13 (26.0) | p=0.21 |
| College or more | 78 (79.6) | 41 (85.4) | 37 (74.0) |  |
| Medication/Vitamin, any |  |  |  |  |
| No | 83 (90.8) | 44 (91.7) | 39 (78.0) | p=0.09 |
| Yes | 15 (9.2) | 4 (8.3) | 11 (22.0) |  |
| Median days (range) between MRI and first intervention. |  | 10 (0-21) | 10 (1-25) | NS |

### Primary Outcomes

#### Clinical Outcomes

The primary outcome was measured as the change from baseline to week 20. Headache frequency was measured using an electronic daily diary for 28 days based on the National Institute of Neurological Disorders and Stroke preventive therapy headache diary, which was provided via an email link. When fewer than the full 28 days were completed, the proportion of headache days was calculated (number of headache days/total number of diary days) and then multiplied by 28 to get a continuous variable for headache days. Note: in the clinical trial registration we included headache-related disability as a primary outcome. However, given prior migraine RCTs have almost exclusively used headache frequency as the primary outcome, we chose to limit focus on that sole primary outcome.

#### Imaging Outcomes

Brain function was measured as activation during cognitive task [15] performance in left DLPFC and cognitive task network (EMN), and resting state connectivity of right dorsal anterior insula to left DLPFC and cognitive task network (EMN). Brain structure was measured as gray matter volume in DLPFC, cingulate, and anterior insula. Regions-of-interest (ROIs) were defined from the cognitive task group activation map for all participants combined at baseline. Peak voxels for each region were selected, a 4mm radius sphere was created, and data were extracted from the scan of interest for each subject. Further details on neuroimaging data and analysis are provided below.

### Secondary outcomes

#### Clinical Outcomes

Secondary outcomes were assessed at weeks 10, 20, and 52. Headache-related disability was measured using the six item Headache Impact Test [43] that shows strong psychometric properties [78]. Headache intensity was computed as the average of all headache intensity ratings from the electronic daily diary [69]. Response to treatment was defined as >50% reduction in number of headache days [69] from baseline to week 20. A migraine [15] day was coded when at least 2 of the following criteria were met: unilateral, pulsating, moderate/severe pain, aggravated by routine activity; and at least one of the following criteria were met: nausea/vomiting or light/noise sensitivity

#### Imaging Outcomes

Whole brain analyses of gray matter volume, activation to pain, activation to cognitive challenge, and resting state connectivity of the insula cortex were measured using Sandwich estimator toolbox [36] (see below).

### Sample size

Using a 0.050 two-sided significance level, a sample of 90 subjects randomized to 2 treatment groups (1:1) provides 80% power to detect an effect size (Cohen’s d) of at least 0.60 in change of headache frequency for MBSR+ relative to SMH using a t-test and the difference between a proportion of responders for MBSR of 0.435 (20/45) and for SMH of 0.150 (7/45) using a Fisher’s exact test.

### Subjects included in imaging analyses

MRI analyses were per protocol. Subjects were excluded from imaging for the following reasons, for each scan type: missing data from baseline; if baseline data is available, missing data from weeks 10 and 20; excessive motion; abnormal anatomy; technical issue (most commonly Medoc Pathway failure); claustrophobia or inability to tolerate thermal stimulation; discontinued treatment; refused MRI (at a follow-up visit).

8 subjects were removed from all MRI analyses because they had usable data for only one or no scan sessions, and 2 others were removed because they did not complete treatment. An additional subject was excluded because of a frontal lobe encephalomalacia due to olfactory meningioma removal (late disclosure). Two other subjects were excluded from fMRI analyses only, one who had very high motion, one had no usable functional data. See Supplementary Table 1 for subjects included in each analysis.

### MRI Data Acquisition

T1 MPRAGE (repetition time (TR) 2300ms, echo time (TE) 2.98ms, slice thickness 1mm, field of view (FOV) 256mm, flip angle 9°, voxel size 1×1×1mm), high resolution anatomical scan for template registration and gray matter volume analysis, a resting state functional MRI scan (10 minutes, echo planar imaging, EPI, TR 2000ms, TE 28ms, slice thickness 4mm, FOV 220mm, flip angle 77°, voxel size 3.4×3.4×4mm), an fMRI scan with blocks of painful thermal stimulation (2 runs of 8 minutes); an fMRI scan with cognitive task (two runs of 5 minutes 10 seconds). Parameters for pain and cognitive task scans: EPI, TR 2500ms, TE 30ms, slice thickness 3mm, FOV 230mm, flip angle 90°, voxel size 3×3×3×mm. A diffusion weighted scan for diffusion tensor imaging (DTI) and a resting state arterial spin labeling (ASL) scan were also collected, but are not analyzed here.

### Voxel-Based Morphometry (VBM)

We used voxel based morphometry (VBM) to assess longitudinal GMV changes in patients as well as to assess differences between patient treatment groups over time [5]. All images were realigned to the anterior-posterior commissure in Statistical Parametric Mapping (SPM12) prior to pre-processing. The computational anatomy (CAT12) toolbox located within SPM12 was used for the longitudinal preprocessing of patient T1 images [31]. Using the longitudinal segmentation pipeline in CAT12.1 (r1278) the structural T1-weighted images acquired at each time point were spatially normalized to Montreal Neurological Institute (MNI) space (resampled to a voxel size of 1.5×1.5×1.5mm), segmented into grey matter (GM), white matter (WM) and cerebrospinal fluid (CSF). Scans were preprocessed with an absolute threshold mask of 0.1. This threshold excluded voxels with less than 10% probability of being grey matter. Finally, images were smoothed with an 8mm Gaussian Kernel prior to analysis.

Quality controls of MRI images took place in two stages. Raw images were carefully examined for morphological abnormalities, as well as movement and scanner artifacts prior to pre-processing. Each image was also overlaid with the T1 template in Check Reg to assess the orientation. Post preprocessing each image was assessed for quality using the two options available in CAT12 [31]. We utilized the display all slices option in CAT12, which displays one horizontal slice for each subject and thus gives an overview of the segmentation. Any images that warranted further inspection were overlaid on the standard T1 brain in Check Reg. Additionally we utilized the sample homogeneity option in CAT12 and the quality measures created during preprocessing as well as nuisance parameters (age, sex and TIV) to get a better picture of the quality of the data. The tool displays a correlation matrix as well as overall mean correlation and weighted overall image quality [31]. The option to view the most deviating data was also used and any images presented in this tool were further examined in Check Reg. Provided data did not have any artifacts upon further inspection they were utilized in the analysis.

### Resting state fMRI

Preprocessing was performed in SPM12 and included: slice timing correction; realignment (motion correction); co-registration of the T1 to the mean functional image; segmentation of the T1; normalization of functional images, with interpolation to 2×2×2mm voxels; smoothing of 6mm. Note that resting state fMRI underwent additional preprocessing. Quality control steps included visual inspection of the data at each preprocessing stage. SPM12 defaults were used except in rare cases were preprocessing led to suboptimal registration or normalization.

We applied a motion regression approach based on framewise displacement using custom scripts. We removed subjects if FrameWise Displacement Arithmetic Mean was greater than 0.3 [52-54]. We chose this cutoff because it appeared to consistently remove subjects whose average motion was larger than the rest of the group. We did not observe any differences in movement across sessions or conditions.

Insula seeds were selected based on the study “Decoding the role of the insula in human cognition: functional parcellation and large-scale reverse inference” [17]. This paper used hierarchical clustering of resting state functional connectivity of insula voxels and achieved a three parcellation solution: dorsal anterior insula, ventral anterior insula, and posterior insula. Based on reverse inferense from neurosynth.org, ventral insula was associated with affective responses, dorsal with cognitive, and posterior with sensory. These regions of interest were downloaded from https://identifiers.org/neurovault.collection:13 following correspondence with the authors. The seeds were masked with the AAL2 atlas region for insula because the original ROIs extended outside of the insula.

Data processing was done via CONN toolbox [77]. We used 5 principal components for white matter (WM) and cerebrospinal fluid (CSF) using triply eroded WM and and eroded CSF. Additionally, we used motion parameters and first order temporal derivative. Applied linear detrending and a bandpass filter of 0.008 to 0.1 Hz (filtering applied simultaneously). Motion spikes were identified using a output from a custom script and included movement ‘spikes’ where greater than .3 mm of framewise displacement occurred.

Subjects underwent first level analysis of resting state functional connectivity for each ROI together. Group-level seed maps (1 way ANOVA), calculated from baseline data (n=74), were as expected (Supplementary Figure 1). Dorsal aINS was strongly linked to attention networks (dorsal attention, salience, DLPFC, EMN). pINS was strongly connected to thalamus and sensorimotor cortex and parts of posterior mid cingulate cortex [72]. Ventral aINS was functionally connected with DMN and amygdala. These findings are consistent with [17].

### Multi-Source Interference Task (MSIT) fMRI

Particpants performed a cognitive task, the multi-source interference task task (MSIT) [15], which reliably activates the mid cingulate cortex and EMN [59], as reported previously [59;66]. Subjects were trained to perform the task prior to the scanning session, then allowed again to practice the session in the scanner before the session began. First-level analyses: 2 runs of 5min 10s, where each run included 5×20s blocks of control, 5×20 blocks difficult, with 10s of tapping in between each task block. Motion parameters were included in the GLM. A one-sample t-test was used to create a baseline group MSIT map comparing the difficult and easy tasks (n=81; Supplementary Figure 2).

### Pain fMRI

Subject-specific,moderately painful (5-7 out 10 on a numeric rating scale) temperatures were selected based on subjects’ responses to pre-scan quantitative sensory testing and confirmed with verbal ratings once patients were inside the scanner. Thermal stimuli were applied to the left forarm using Medoc Inc Pathway with ATS 30mm thermode. Two runs of 8min, where each run included 5 stimuli of 28s ramp and hold at a nonpainful warm temperature and then at the subject-specific, moderately painful temperature. After each run, subjects verbally rated the average and maximum pain intensity on a 0-10 numeric scale. First-level analyses: warm and pain onset were modeled as the first 2s of the stimulus, while warm and pain block were modeled as the subsequent 28s. Additionally, pain offset was modeled as the 2s starting at the onset of descending ramp back to the 32°C (baseline). Motion parameters were included in the GLM. One-sample t-tests are shown for baseline (n=77) pain onset (Supplementary Figure 3) and pain block (Supplementary Figure 4).

### Statistical Methods

Clinical outcomes were analyzed using the intention-to-treat approach. Effects of intervention were estimated using mixed-effects models, where patient was a random effect, and fixed factors included treatment, time, treatment-by-time interaction, age, cohort, interval in days between MRI and treatment, medication, presence of other pain, and education. Difference in treatment response rate was assessed using a generalized linear model with a logit link function. The regression model for the mean with the binomial distribution variance function was utilized to model the log odds ratios. The generalized linear models included the following covariates: age, medication, level of education, presence of other pain, interval in days between MRI and first intervention. A logistic regression model predicted probability of response to treatment. P-values are nominal and not adjusted for multiple outcomes. Testing was two-sided and used the 0.05 level of significance, and statistical analyses used R-Studio, Version 1.1.453.

Imaging outcomes were analyzed per protocol.

### Regions of Interest (ROIs)

Our primary outcomes for MRI included: left DLPFC and EMN activation during cognitive task performance; DLPFC, cingulate, and aINS GMV; resting state connectivity of the right dorsal aINS to the left DLPFC and component regions of the EMN. ROI selection was based on hypothesized areas that are involved in both pain and cognition [59]. We used the peak voxels from group-level (n=81) MSIT activation and created a 4mm sphere around each peak. The regions selected included the left and right DLPFC, left and right aINS, and aMCC. We also created a mask for all MSIT activations (EMN) [29].

ROI analyses used the same linear mixed models approach as used for primary clinical outcomes. Linear mixed models included patient as a random effect and fixed factors of treatment, time, and the treatment-by-time interaction, and scanner as a covariate, with bonferroni correction for multiple comparisons.

### Whole Brain analyses

#### Sandwich Estimator Toolbox Analysis (SwE)

The Sandwich estimator toolbox (SwE) (version 2.0) was used to model the longitudinal changes in GMV in patients [36]. This toolbox is specifically designed for repeated measures MRI analysis and uses an unstructured covariance structure and a small sample adjustment. It uses an alternative to the traditional linear mixed model and employs a simple Ordinary Least Squares (OLS) marginal model for estimates of the parameters of interest. The sandwich estimator is utilized to calculate the standard errors of these estimates and is used in conjunction with the OLS. The flexibility of the SwE is in its robustness to misspecification of the covariance structure and the utilization of this approach accounts for the correlations in repeated measures and can be used with unbalanced datasets with missing data.

We modeled treatment group (MBSR vs. SMH) by time (baseline, week 10, week 20) interactions to compare change in GMV over time between the two treatments. Additionally, to account for the scanner upgrade which occurred within the last year of the study and impacted the last of the 8 cohorts of patients enrolled (patients with scans conducted on both scanners), all analyses were run adjusting for this change. Secondary analysis examined the treatment and time effects.

Using the non-parametric SwE model with 10,000 bootstraps, we used a cluster-forming threshold of p<0.001 and FWE (estimated from the wild bootstrap distribution) correction of 0.05 at the cluster level. The contrast of interest was the treatment-by-time interaction. Main effects of time and treatment are provided in Supplementary.

## Results

Among 573 individuals contacted for telephone screening, 168 were potentially eligible and 119 of these met the headache frequency criteria during baseline (Figure 1). The main reasons for exclusion were not meeting migraine or headache frequency criteria, ineligible or refused MRI/pain testing, schedule incompatability, or migraine secondary to injury. Ninety eight participants were randomized to treatment; 50 were assigned to receive MBSR+ and 48 were assigned to receive SMH. All attended at least one session. Five participants (5%) withdrew from treatment after the first session but agreed to continue with data collection and 3 participants (3%) withdrew from treatment and were lost to follow-up. Forty three (86%) of the MBSR+ participants and 40 (83%) of the SMH participants completed all sessions, either in the group or individually as a make-up.

At baseline, treatment groups were similar on all sociodemographic characteristics (Table 1). Participants (mean age of 36 years) were predominantly female (91%), white (72%), and 80% had completed at least one year of college. At baseline, they reported an average of 7.8 headache days and only 15% were using a preventive treatment for migraine. There were no group differences in treatment withdrawal or loss to follow-up.

### Primary outcomes

At week 20, the MBSR+ group reported fewer headache days (4.6 [95% CI 3.6 to 5.6]) compared to the SMH group (6.0 [95% CI 4.9 to 7.0]; *P*=.04; Table 2). This effect was apparent at week 10, as the MBSR+ group reported fewer headache days (5.5 [95% CI 4.6 to 6.5]) compared to the SMH group (6.9 [95% CI 5.9 to 7.9] *P*=.04). This treatment effect was not significant at week 52 (*P*=.12).

**Table 2.** Primary and Secondary Clinical Outcomes. Adjusted means, 95% Confidence Intervals.

| **Primary Clinical Outcomes<sup>1</sup>** |  |  |  |
| --- | --- | --- | --- |
| Week | SMH | MBSR+ | p-value |
| **Headache days (per 28 day calendar)<sup>2</sup>** |  |  |  |
| baseline | 7.7 (6.7 8.7) | 7.8 (6.9 8.8) | 0.85 |
| 10 | 6.9 (5.9 7.9) | 5.5 (4.6 6.5) | **0.04** |
| 20 | 6.0 (4.9 7.0) | 4.6 (3.7 5.6) | **0.04** |
| 52 | 5.6 (4.6 6.7) | 4.6 (3.7 5.6) | 0.12 |
| **Secondary Clinical Outcomes** |  |  |  |
| **HIT-6<sup>3</sup>** |  |  |  |
| baseline | 59.6 (57.7 61.5) | 59.6 (57.9 61.3) | 0.99 |
| 10 | 58.5 (56.5 60.4) | 56.3 (54.5 58.1) | 0.08 |
| 20 | 57.5 (55.5 59.4) | 54.6 (52.9 56.4) | **0.02** |
| 52 | 58.4 (56.4 60.4) | 56.2 (54.4 58.1) | 0.10 |
| **Pain severity<sup>4</sup>** |  |  |  |
| baseline | 4.3 (3.8 4.8) | 4.7 (4.2 5.2) | 0.20 |
| 10 | 4.4 (3.9 4.9) | 4.4 (4.0 4.9) | 0.62 |
| 20 | 4.4 (3.9 5.0) | 4.4 (4.0 4.9) | 0.63 |
| 52 | 4.7 (4.2 5.3) | 4.5 (4.1 5.0) | 0.84 |
| **Number of responders<sup>5</sup> N, %; 95% CI** |  |  |  |
| 20 | 11 (23%; 12-37%) | 26 (52%; 37-66%) | **0.004** |
| **Migraine days (per 28 day calendar)<sup>2</sup>** |  |  |  |
| baseline | 3.2 (2.2 4.2) | 3.3 (2.5 4.2) | 0.83 |
| 10 | 4.0 (3.0 5.0) | 1.9 (1.0 2.8) | **0.0008** |
| 20 | 3.7 (2.7 4.7) | 2.0 (1.1 2.9) | **0.004** |
| 52 | 3.1 (2.1 4.0) | 2.1 (1.2 3.0) | 0.12 |

<sup>1</sup>Primary clinical outcome time point was week 20 (gray shading).
<sup>2</sup>adjusted for cohort, age, medication, and education level.
<sup>3</sup>adjusted for cohort, age, medication, and presence of idiopathic pain.
<sup>4</sup>adjusted for cohort, age, medications, education level, presence of idiopathic pain, and interval days between baseline and first MBSR/SMH session.
<sup>5</sup>Response was defined as 50% or greater reduction in headache frequency at week 20 compared to baseline.

Regions of interest revealed no significant treatment-by-time effects related to gray matter volume, cognitive task activation, or resting state fMRI (see Supplementary). Both groups showed decreased anterior mid cingulate volume (P=.04) and decreased connectivity of right dorsal anterior insula to cognitive task network (EMN) (P=.02) at week 20.

### Secondary outcomes

At week 20, the MBSR+ group also reported reduced HIT-6 scores (2.0 [95% CI 1.1 to 2.9]) compared to the SMH group (3.7 [95% CI 2.7 to 4.7]; *P*=.04). Headache impact did not differ between treatment groups at week 10 or week 52 (Table 2) and average headache pain intensity did not differ between treatment group at any timepoint (Table 2). At week 20, 52% of the MBSR+ group were classified as treatment responders (>50% reduction in headache days) compared to 23% of the SMH group (*P*=0.004; Table 2, Figure 2), yielding a number needed to treat (NNT) of 3.4. The MBSR+ group reported fewer migraine days at week 10 (*P*=.0008) and week 20 (*P*=.004) relative to SMH, but not at week 52 (Table 2).

**Figure 2.**
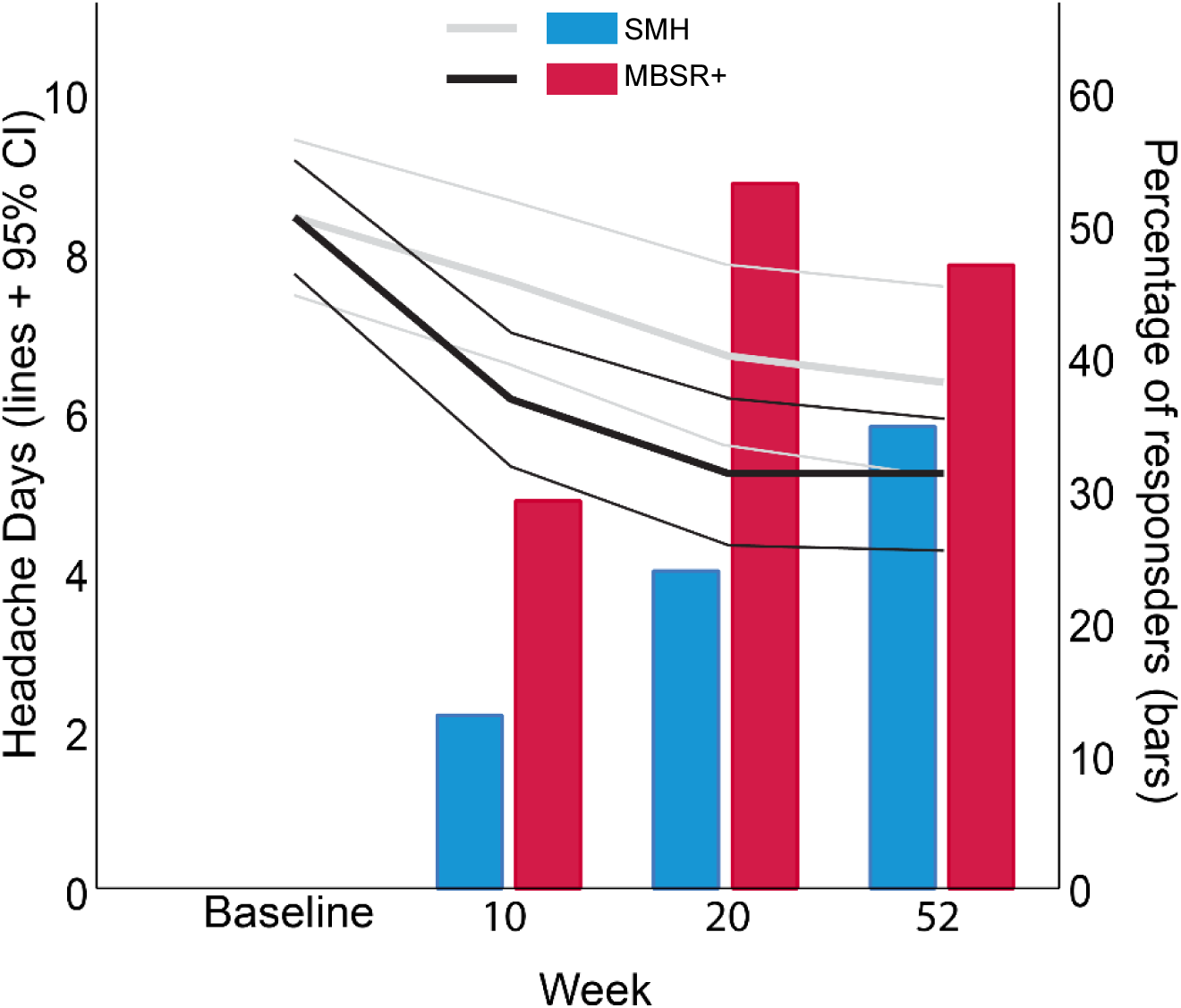
Clinical outcomes. Mean (thick lines; 95% confidence intervals shown in thin lines) number of headache days per 28-day diary (uncorrected values) for SMH (gray) and MBSR+ (black). Responder rates (with response defined as a 50% reduction in headach days from baseline) are shown in bar plots, with MBSR+ in red and SMH in blue. At week 20 (primary outcome) both headache frequency and response rate were significantly better in the MBSR+ group.

Whole brain analyses revealed a significant treatment-by-time interaction on activation during the cognitive challenge. The MBSR+ group showed decreased activation in the bilateral cuneus and right parietal operculum at week 20 compared to the SMH group (Figure 3; Supplementary Table 2). Whole brain analyses also revealed a significant interaction of left dorsal anterior insula connectivity to the right posterior parietal cortex and right cuneus (Figure 3; Supplementary Table 2). There were no significant interaction effects for the other five insula seed regions, gray matter volume, or activation during pain stimulation for the whole brain analyses.

**Figure 3.**
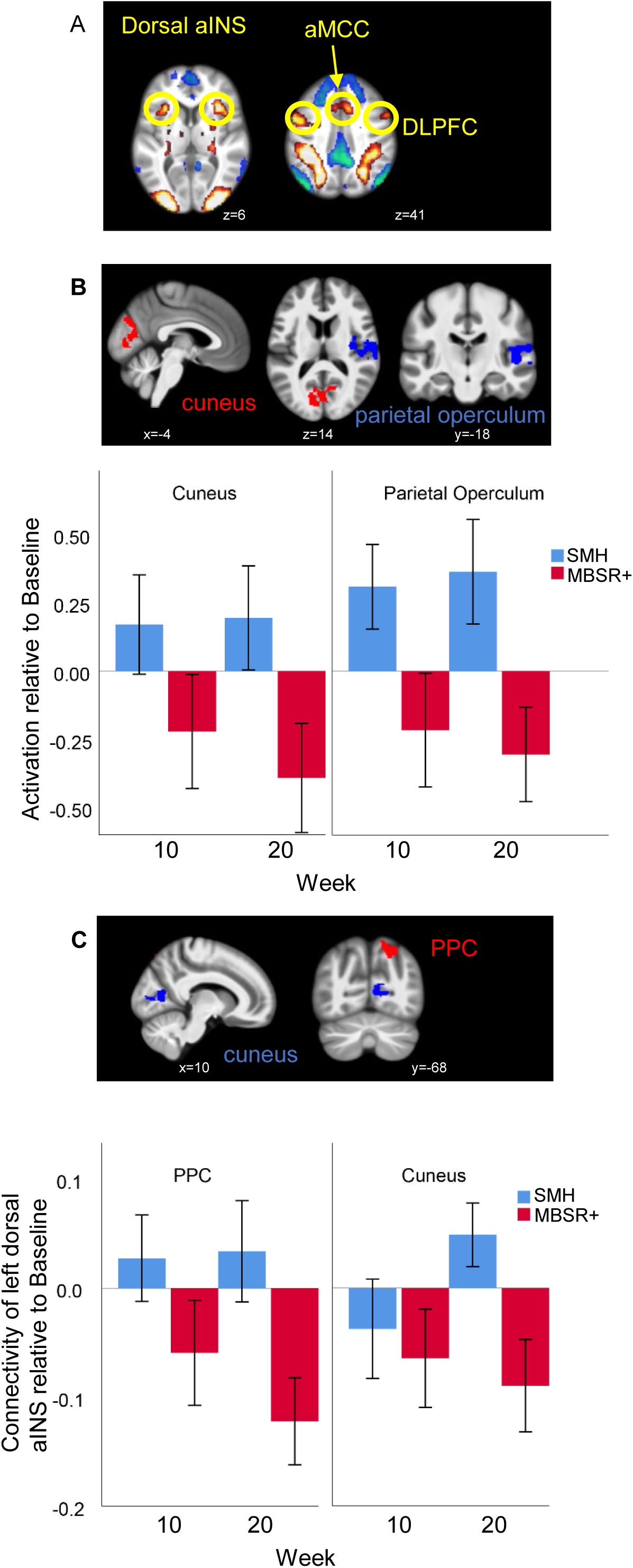
MRI outcomes. A) Regions-of-interest for primary outcome analyses. The activation map is defined from the baseline the baseline group map (all patients) for activation during the cognitive task (difficult vs easy contrast on the multi-source interference task). The circled regions show the five ROIs derived from this map (left and right dorsal anterior insula (aINS), left and right dorsolateral prefrontal cortex (DLPFC), and anterior midcingulate cortex (aMCC). These ROIs were used to assess the primary MRI outcomes, which included structure, resting state connectivity, and pain- and cognitive-related activation. There were no significant effects of treatment for region-of-interest analyses. B) Whole-brain (secondary outcome) analysis interaction effect for cognitive task-related activation. The SwE model assess the interaction between treatment and time for the difficult vs easy contrast on the multi-source interference task. Identified regions show a decrease in activation level in the MBSR+ compared to SMH group over time. C) Whole-brain analysis interaction effect for left dorsal aINS connectivity. Regions show a decrease in activation level in the MBSR+ compared to SMH group over time. Images are displayed on the average anatomical MRI for all patients at baseline. Error bars are 95% confidence intervals. MNI coordinates are shown for each slice as x, y, or z.

### Adverse Events

There were 16 adverse events reported of which 15 were mild (e.g., high blood pressure, hives, jaw pain) or moderate (e.g., car accident, kidney stone); the one serious adverse event (stroke), in accordance with the data safety monitoring plan and consultation with the independent monitoring committee, was deemed unlikely related to intervention. The remaining 7 were definitely not related, 7 were unlikely related, and 1 was possibly related to study procedures (one participant reported a migraine during the MRI session).

## Discussion

Among adults with episodic migraine, enhanced mindfulness based stress reduction (MBSR+) decreased headache and migraine days and headache related disability, as well as yielded a higher treatment response rate, relative to the active control (SMH). Treatment response (50% reduction in headache frequency) to MBSR+ relative to SMH yielded an NNT of 3.4, which is comparable to valproic acid – one of the first line treatments for episodic migraine prophylaxis [45]. These results hold promise for the use of mindfulness-based interventions for headache, with treatment response rates qualitatively comparable or exceeding effects of most existing standard pharmaceutical therapies in the time frames they have been tested [2; 6; 7; 10; 12; 24-26; 30; 46; 47; 56; 67; 68; 70].

While the effects of MBSR+ in reducing headache frequency were not significantly different from SMH at the 52 week follow-up, it is worth noting that the reduced headache frequency of 4.6 headaches in the MBSR+ group observed at post-treatment remained steady through to 52 weeks (see Table 2). What changed at 52 weeks was the headache frequency of the SMH group, which showed a slow, steady reduction in headache frequency over the course of the study, so that the difference between groups became non-significant at 52 weeks. A common issue in conducting clinical trials is the challenge of “regression to the mean,” since study participants may be experiencing a particularly difficult period with symptoms at the time of enrollment, and the design solution is inclusion of a control group. We designed the study to include an active control group, led by an experienced nurse expert in headache management to account for the influence of expectations and non-specific effects of intervention as well as the effects of time. This pattern of findings suggests that the SMH intervention, which included discussions of multiple headache management skills (building social support networks and managing diet, exercise, and sleep), may slowly reduce headache frequency.Although no effects of MBSR+ training were observed on the primary neuroimaging outcomes, secondary whole-brain analyses identified two findings that suggest an increase in cognitive efficiency that is consistent with findings from the meditation literature. Compared to SMH, MBSR+ training led to decreased activation of the parietal operculum and visual cortex (cuneus) during the cognitive challenge. Both long-term meditators [9] and individuals trained in MBSR [41] show altered visual cortex connectivity and increased activation during focused attention. The parietal operculum, including posterior insula, is activated by pain and deactivated by cognitive challenge [59] and we have previously reported that this is the only acute pain-related activation that is modulated by cognitive demand in both healthy subjects and migraine patients [48]. Additionally, we observed reduced resting connectivity of the dorsal anterior insula to posterior parietal cortex and visual cortex (cuneus) following MBSR+ training compared to SMH. Because dorsal anterior insula strongly connects to the posterior parietal cortex and cuneus as part of the cognitive task network [17], this finding supports increased cognitive efficiency following MBSR+. These increases in cognitive efficiency seen in the MBSR+ group may reflect changes due to the practice of meditation or alternatively may reflect the effect of having fewer headaches during the period surrounding measurement.

Our study examined MRI primary outcomes in a registered clinical trial for a chronic pain condition. The primary imaging outcomes, including changes in gray matter volume, activation during cognitive challenge, and resting state connectivity of the anterior insula in *a priori* selected regions-of-interest, did not differ between groups. The choice of these regions was based on literature when the study was proposed and our preliminary data, focusing on the DLPFC [63] and other brain areas showing pain-cognition interactions [16; 48; 60; 62; 64; 66]. The vast majority of neuroimaging studies compare individuals with chronic pain to healthy subjects, rather than longitudinal designs examining how the brain changes with treatment. Since we did not find treatment effects in the areas that distinguish those experiencing daily pain, our findings suggest that brain changes distinguishing patients from healthy controls might not be useful as treatment targets.

Most MRI studies reporting effects of treatment have only investigated the treatment group [37; 55; 65; 66] or treatment responders within a group exposed to treatment [27; 73; 74]. This study compares two active treatment arms and includes both treatment responsders and nonresponders in the analyses. It is possible that the results from previous studies examining brain changes over time are dependent on treatment response, rather than the effects of the specific intervention itself. Future work should thus include comparisons of responders and non-responders, as well as examining the associations between changes in clinical and neuroimaging outcomes.

These findings share limitations common to most RCTs and may have limited generalizability due to the likely selection bias that results from the strenuous requirements of participation, including time commitment and willingness to complete repeated MRI scans, resulting in most of the participants being college educated. Study strengths, in addition to the use of MRI outcomes, include one of the largest sample sizes for measuring brain imaging outcomes in migraine or any chronic pain disorder, the very small loss to follow-up, the close matching of MBSR+ to the active control, and long-term follow-up.

## Conclusions

In episodic migraine, MBSR+ showed superior treatment effects compared to an active control, with significant reductions in headache frequency that are comparable to commonly used first line treatments for episodic migraine prophylaxis. Brain changes in the MBSR+ group were seen in the pattern of functional connectivity and activation during a challenging cognitive task that are consistent with increased cognitive efficiency. These findings suggest that MBSR+ can be an effective prophylactic treatment option for episodic migraine.

## Data Availability

Available upon request from corresponding author

## Data Availability

Available upon request from corresponding author

## Acknowledgements

We thank the Independent Monitoring Committee members: Erica Sibinga, Tianjing Li, George Wittenberg, and Michael Baime. The authors declare no conflicts of interest.

